# Efficacy and Safety of Middle Meningeal Artery Embolization in Chronic Subdural Hematoma: A Comprehensive Systematic Review and Meta-Analysis

**DOI:** 10.64898/2025.12.31.25343272

**Authors:** Farzan Fahim, Negin Safari Dehnavi, MohammadAmin Farajzadeh, Aysan Valinejad, Shahriar Heshmaty, Amirhossein Rastegar, Zahra Aghabeygi, Fatemeh Begmaz, Aida mahmoudjanlu, Somayeh Golmohammadi, Saeed Oraee-Yazdani, Alireza Zali, Sayeh oveisi

**Affiliations:** Neurosurgery resident, Shohada-E-Tajrish hospital, Shahid Beheshti University of Medical Science, Tehran Iran; Functional Neurosurgery Research Center, Research Institute of Functional Neurosurgery, Shohada Tajrish Comprehensive Neurosurgical Center of Excellence, Shahid Beheshti University of Medical Sciences, Tehran, Iran; School of Medicine, Shahid Beheshti University of Medical Sciences, Tehran, Iran; Associated professor of neurosurgery, functional neurosurgery of research center, Shohada-E-Tajrish hospital, Shahid Beheshti University of Medical Science, stereotactic fellowship, Tehran, Iran; Professor of neurosurgery, functional neurosurgery of research center, Shohada-E-Tajrish hospital, Shahid Beheshti University of Medical Science, stereotactic fellowship, Tehran, Iran

**Keywords:** Middle meningeal artery embolization, Chronic subdural hematoma, Recurrence reduction, Procedural safety, Endovascular therapy, Meta-analysis

## Abstract

**Background:** Middle meningeal artery embolization (MMAE) has emerged as an adjunct or alternative strategy for the management of chronic subdural hematoma (cSDH). Although accumulating studies suggest potential benefit, uncertainty remains regarding its safety profile, recurrence-prevention effect, and the reliability of adverse event reporting. This systematic review and meta-analysis re-evaluate contemporary evidence, incorporating new randomized trials and large observational cohorts.

**Methods:** This systematic review was conducted in accordance with PRISMA 2020 guidelines and prospectively registered in PROSPERO. PubMed, Scopus, Web of Science Core Collection, Embase, and CENTRAL were searched from inception to 12 September 2025 without language restrictions. Randomized controlled trials, prospective or retrospective cohort studies, and non-randomized clinical studies evaluating middle meningeal artery embolization (MMAE) for chronic subdural hematoma were eligible. Data extraction and risk-of-bias assessment were performed independently using Joanna Briggs Institute appraisal tools. Where outcomes were sufficiently comparable, quantitative synthesis was undertaken using random-effects single-arm proportion meta-analysis with logit transformation. Recurrence after MMAE was pooled across observational studies and MMAE arms of randomized trials with available event-level data, with prespecified subgroup analyses by study design. Mortality was synthesized from randomized trials reporting event-level data within a ≤90-day follow-up window. Complication rates and technical success were analyzed descriptively due to heterogeneity in definitions and follow-up durations.

**Results:** Nineteen studies met eligibility criteria, including seven randomized controlled trials, sixteen retrospective cohorts, and one prospective cohort, comprising an elderly and medically complex population (mean ages 61–89 years). Common comorbidities included hypertension, diabetes, cardiovascular and cerebrovascular disease, renal dysfunction, and antithrombotic use. Technical success of middle meningeal artery embolization (MMAE) was consistently high, with a pooled success rate of 100% (95% CI 0.99–1.00; I^2^ = 0%). Recurrence after MMAE was consistently low across randomized and observational studies, including high-risk populations, and was uniformly lower than in comparator groups. Radiographic outcomes showed substantial hematoma volume reduction and high rates of complete or near-complete resolution, with favorable functional recovery. Complications were uncommon but heterogeneous; the pooled overall complication rate was 14% (95% CI 0.08–0.21). Pooled 90-day all-cause mortality from randomized trials was 8% (95% CI 0.07–0.10; I^2^ = 0%).

**Conclusion:** MMAE is a safe and effective adjunctive or alternative treatment for chronic subdural hematoma, demonstrating a reproducible and clinically meaningful reduction in recurrence across randomized and observational datasets with homogeneous outcome definitions. However, variability in adverse event reporting, insufficient documentation of rare complications, and inconsistent definitions of radiographic versus clinical recurrence highlight the need for standardized outcome frameworks and harmonized follow-up protocols. Future well-designed trials with robust adverse event adjudication are essential to define the long-term safety profile of MMAE and to guide its optimal integration into cSDH management pathways.

## Introduction

Chronic subdural hematoma (cSDH) is generally defined as a slowly accumulating collection of blood and fluid on the brain surface, located between the dura mater and the arachnoid membrane^1^. cSDH occurs predominantly in older men; head trauma and chronic alcohol consumption are frequent etiological factors in this group^2^. More recent experimental and clinical work indicates that persistent inflammatory activity within the hematoma’s membranes plays a central role in its development and progression.

From a service perspective, cSDH is one of the most frequent reasons for neurosurgical consultation in older adults, and its incidence continues to increase as populations age^1^. Reported annual incidence in the general population is approximately 8–14 cases per 100,000 people, rising to about 60–70 cases per 100,000 among elderly individuals^3^. Clinical presentation varies; patients may present with motor deficit or reduced level of consciousness, and these symptoms often lead to substantial functional decline if treatment is delayed^2^.

Standard management is surgical evacuation of the hematoma, most often through burr-hole craniotomy. This approach is effective in many cases, yet recurrence still occurs in about 12% of patients^3^. Surgery itself may result in complications such as seizures and infections, and frail patients can experience further deterioration in functional status after the procedure^4^. In response to these problems, several pharmacological regimens including corticosteroids, statins, and tranexamic acid have been evaluated as adjunctive or alternative treatments aimed at lowering recurrence and supporting hematoma resolution^5^.

Middle meningeal artery embolization (MMAE) has been introduced as a minimally invasive endovascular option for cSDH and was first used around 2000 as a salvage strategy in selected patients^6^. By occluding branches of the middle meningeal artery that supply the vascularized outer membrane, MMAE targets the presumed source of repeated microbleeding and inflammatory drive within the dura^7^. The intention is to modify the underlying biological process and to decrease the likelihood of hematoma regrowth.

Several clinical series and randomized controlled trials now suggest that MMAE can reduce the recurrence of cSDH compared with conventional management alone^8,9^. Many of these studies, however, report follow-up of only about 90–180 days, which limits the assessment of later outcomes^10,11^. In addition, evidence regarding functional recovery, rate of hematoma volume reduction, appropriate patient selection, and mortality remains incomplete or inconsistent across reports^12^. Some investigators have proposed MMAE as a standalone treatment, whereas others have reported better results when it is combined with surgical evacuation, creating uncertainty in clinicians’ routine decision-making^13^.

Initial aggregated data are promising for both efficacy and safety of MMAE. Yet, the current literature still does not provide a definitive answer on long-term benefit or on uncommon but serious complications^14^. New randomized controlled trials and updated meta-analyses have appeared in the last few years, so there is a clear need to re-evaluate the safety profile of MMAE in cSDH using contemporary evidence^15^. This systematic review and meta-analysis was therefore designed to examine the safety of MMAE in patients with chronic subdural hematoma, with a specific focus on the incidence and severity of adverse events associated with the procedure.

## Method

### Registration, protocol, and reporting

This systematic review was planned and reported in accordance with the PRISMA 2020 statement. The protocol was registered prospectively in PROSPERO (CRD420251251568). The PRISMA checklist, the verbatim search strategies for all databases, and the final data extraction form are available in the supplementary materials.

### Search strategy and information sources

A comprehensive search of electronic databases was conducted in PubMed, Scopus, Web of Science Core Collection, the Cochrane Central Register of Controlled Trials, and Embase. Each database was searched from inception to 12 September 2025. No date limits were imposed, and we didn’t apply any language restrictions at the search stage.

Search strategies combined controlled vocabulary (for example, MeSH in PubMed and Emtree in Embase) with free-text terms capturing three main concepts: the middle meningeal artery or transarterial procedures; embolization or embolotherapy; and chronic or refractory subdural hematoma or hemorrhage. Appropriate Boolean operators, truncation, and field tags were used according to the syntax of each database. The exact line-by-line strategies for all platforms are reproduced verbatim in the supplementary material, including spelling, spacing, and operators as implemented.(supplementary 1)

### Eligibility criteria

Eligibility criteria were defined a priori using a PICO(S) framework. The population of interest comprised adult patients diagnosed with chronic subdural hematoma. The intervention was middle meningeal artery embolization, regardless of embolic material, unilateral or bilateral treatment, or technical details such as catheter system or target branch. Comparator groups included any management pathway in which embolization of the middle meningeal artery was not performed, for example surgical evacuation alone, medical management, or other interventional strategies.

The primary outcomes were safety related, with a focus on procedure-related complications, recurrence of chronic subdural hematoma after treatment, and mortality. Secondary outcomes were related to treatment effect, including change in hematoma volume and functional recovery as reported by the original studies. Eligible designs were randomized controlled trials, prospective cohort studies, retrospective cohort studies, and non-randomized clinical trials.

Studies were excluded if they were single-patient case reports, animal or in vitro work, narrative reviews, letters or commentaries, conference abstracts without sufficient data, duplicate publications, or if they did not provide adequate information on safety or efficacy outcomes associated with middle meningeal artery embolization.

### Study selection

All records retrieved from the database searches were imported into EndNote, where automated and manual procedures were used to identify and remove duplicates. The de-duplicated dataset was exported into Microsoft Excel for screening.

Screening was conducted in two stages. In the first stage, titles and abstracts were independently evaluated by three reviewers (AM, SH, AR) against predefined eligibility criteria. Any disagreements in this step were resolved by discussion with a fourth reviewer (NSD). Studies that appeared to meet the criteria, or where relevance was uncertain, were taken forward to full-text review. In the second stage, the full texts were independently assessed by two reviewers (AV and AR). Conflicts were resolved through consultation with a third reviewer (NS) until consensus was reached. Reasons for exclusion at the full-text stage were recorded systematically. A complete list of included and excluded studies, with reasons for exclusion, is presented in the supplementary material.(supplementary 2)

### Data extraction

A standardized data extraction form was developed in Microsoft Excel by FF before data collection. The form covered general study characteristics such as first author, year of publication, country, study design, and total sample size; patient characteristics including age, sex, comorbidities, and use of antithrombotic or anticoagulant agents; detailed information on the embolization procedure, including type, size and dose of embolic material, commercial product, unilateral or bilateral middle meningeal artery embolization, timing of the intervention in relation to diagnosis or surgery, target branch (proximal or distal), catheter and microguidewire specifications, use of a guiding sheath, and type of anesthesia; characteristics of the comparator group; and reported safety and outcome measures relevant to the review question.

One reviewer (SH) extracted data from all included studies using this template. A second reviewer (NSD) checked the extracted data for accuracy and internal consistency. Any discrepancies or missing fields identified during this process were resolved by discussion, and corrections were made before the dataset was finalized for analysis.(data extraction sheet is available in supplementary 3)

### Risk of bias assessment

The methodological quality and risk of bias of the included studies were assessed using the Joanna Briggs Institute (JBI) critical appraisal tools, selected based on the study design (cohort and non-randomized clinical studies). Two reviewers (AV and SH) applied the appropriate JBI checklist independently to each study. When differences in judgement occurred, these were discussed with a third reviewer (NSD) until agreement was reached.

The JBI tools include items addressing clarity of participant selection, handling of potential confounding factors, reliability of outcome measurement, completeness of follow-up, and suitability of the statistical methods used. Item-level ratings and overall risk-of-bias judgements for each study are reported in the result.

### Data synthesis and statistical analysis

Extracted data were first summarized descriptively, with tabulation of study design, baseline characteristics of the included populations, details of middle meningeal artery embolization and comparator interventions, and the main reported safety and efficacy outcomes. Where at least two studies reported outcomes that were sufficiently similar in definition and measurement, quantitative synthesis was undertaken.

Recurrence after MMAE was synthesized using a single arm proportion meta analysis including both observational cohorts and the MMAE arms of randomized controlled trials. Event level recurrence data were available for six studies. Pooled recurrence proportions were estimated using a random effects model with logit transformation (metaprop function). Prespecified subgroup analyses were conducted according to study design (RCT vs cohort).

All cause mortality after MMAE was synthesized using a single arm proportion meta analysis restricted to randomized controlled trials reporting event level mortality within a harmonized ≤90 day follow up window. Pooled mortality proportions were estimated using a random effects model with logit transformation (metaprop function). Formal assessment of publication bias was not performed due to the small number of contributing studies.

Overall complication rates following MMAE were explored using a descriptive single arm proportion meta analysis. Studies reporting event level or explicitly stated absence of complications in the MMAE arm were included. Because definitions of complications and follow up durations varied substantially across studies, this analysis was considered exploratory and intended to provide an overall safety signal rather than comparative inference.

Technical success of MMA embolization was optionally explored using a descriptive single arm proportion meta analysis. Technical success was defined as successful catheterization of the target middle meningeal artery with delivery of embolic material as intended, without immediate technical failure or abandonment of the procedure. Because this outcome reflects procedural feasibility rather than clinical efficacy, the analysis was considered descriptive.

Potential publication bias was explored visually using funnel plots and, when the number of contributing studies permitted, by applying the Egger regression test. Sensitivity analyses were conducted by repeating the meta-analyses after exclusion of studies judged to be at high risk of bias. All statistical analyses were performed using Review Manager (RevMan) and Comprehensive Meta-Analysis (CMA) software, with statistical significance defined as p < 0.05.

## Result

### Study characteristics

This systematic review included nineteen studies which published between 2017 and 2025. Studies included seven randomized controlled trials (RCTs), sixteen retrospective cohort studies, and one prospective cohort study ^16^.one study incorporate larger dataset than the others (13,390 patients, of whom 595 received MMAE and 12,795 controls)^12^

The reported mean ages in the included studies ranged from 61 to 89 years, with most of them was between the late 60s and mid-80s. The youngest mean age was 68.2 ± 11.1 years (Luca H. Debs, 2024), and the oldest was 82 years (interquartile range 75–87)^16^

Hypertension was the most prevalent reported comorbidity, followed by Diabetes Mellitus. Cardiovascular illnesses—including Coronary Artery Disease, Myocardial Infarction, and Heart Failure—were frequently described^8,16–18^. Histories of stroke (CVA) were also common across studies ^16,17,19,20^.Other than hypertension, Diabetes and Cardio-Cerebrovascular conditions multiple studies reported renal dysfunction, with Chronic Kidney Disease^16,20,21^. There was also Conditions involving impaired hemostasis—including coagulopathy or thrombocytopenia reported in five of the studies ^8,15,16,20,22^. Additional high-risk factors such as liver dysfunction, malignancy, and chronic alcoholism appeared in selected studies, underscoring the complexity and vulnerability of this patient population. Most studies also reported the use of antiplatelet or anticoagulant therapy, with aspirin and warfarin being the most common agents administered during the pre-procedural period.

**Table 1.** Study Characteristics Table.

| Study | Study Information | Baseline Demographics | Main Medications |
| --- | --- | --- | --- |
| Perng, 2024 | 2024, Taiwan, Retrospective cohort, n=112 | Age: NR; HTN 77.8% | Clopidogrel, Aspirin |
| Schmolling, 2025 | 2025, Spain, Prospective cohort, n=101 | Age: 82 (75–87); HTN 73.5% | AT–APT |
| Dayong Xia,2025 | 2025, China, Retrospective cohort, n=82 | Age: NR; vascular sclerosis & brain atrophy; Male % NR | Oral anticoagulants |
| Eimad Shotar,2025 | 2025, France, RCT, n=319 | Age: 77 (68–83); HTN 59.1% | Anticoagulant / Antiplatelet therapy |
| Martinez-Gutierrez JC, 2024 | 2024, US, Retrospective cohort, n=129 | Age: 74 (65–80); Comorbidities NR | Antiplatelet therapy |
| Catapano, 2021 | 2021, US, Retrospective cohort, n=231 | Age: NR; HTN 57% | Anticoagulant / Antiplatelet therapy |
| Alexander Lam,2023 | 2023, Australia, RCT, n=36 | Age: NR;<br>Coagulopathy<br>12.5% | Aspirin,<br>Aspirin+Ticagrelor,<br>Warfarin |
| Steven B Housley,<br>2023 | 2023, US, Retrospective cohort,<br>n=96 | Age: NR; HTN<br>65.9% | NR |
| J. Liu, 2024 | 2024, China, RCT, n=722 | Age: 69 (IQR<br>61–74); HTN<br>45% | NR |
| Seung Pil Ban, 2017 | 2017, Korea, Retrospective<br>cohort, n=541 | Age: NR; HTN<br>62.5% | NR |
| Christina Onyinzor,<br>2021 | 2021, Germany, Retrospective<br>cohort, n=132 | Age: NR;<br>Comorbidities<br>NR | Antiplatelet /<br>Anticoagulant<br>therapy |
| Luciano Bambini<br>Manzato, 2025, 2025 | 2025, Germany, RCT, n=56 | Age: NR;<br>Diabetes<br>mellitus 21.2% | ASA, Clopidogrel,<br>Apixaban |
| sam ng, 2019 | 2019, France, RCT, n=48 | Age: NR;<br>Trauma history<br>63% | NR |
| Wen Cheng, 2024 | 2024, China, Retrospective<br>cohort, n=112 | Age: 71.94 ±<br>14.35;<br>Underlying<br>diseases 32% | NR |
| Luca H. Debs, 2024 | 2024, US, RCT, n=35 | Age: 68.2 ±<br>11.1;<br>Comorbidities<br>NR | Anticoagulation<br>therapy |
| J.M. Davies, 2024 | 2024, US, RCT, n=400 | Age: 72.0 ±<br>11.2; Baseline<br>similar to<br>control | Similar to baseline |
| El Kim., 2017 | 2017, Korea, Retrospective<br>cohort, n=43 | Age: NR; HTN<br>65% | Antiplatelet /<br>Anticoagulation<br>therapy |
| Adrian Liebert,2023 | 2023, Germany, Retrospective<br>cohort, n=42 | Age: 77.5 ±<br>11;<br>Comorbidities<br>NR | Antithrombotics,<br>Statins |
| Alis J. Dicipinigitis,<br>2025 | 2025, US, Retrospective cohort,<br>n=13390 | Age: NR;<br>Comorbidities<br>NR | Chronic AC/AP use |

### Procedural Details

The procedural approach to Middle Meningeal Artery were different across the studies, the indication for the MMAE in the stdies were mainly; primary treatment of chronic subdural hematoma (cSDH), adjunct therapy following burr-hole drainage (BHD), and standalone treatment for recurrent or non-septated cSDH. Adjuvant use was the most frequent indication, reported in twelve studies. Standalone MMAE was employed multiple studies^8,18,19,21,23^, while adjunctive treatment was performed at various time points relative to surgery. timing of the embolization were varied across studies while J. Liu and colleagues performed MMAE pre-operatively in 99.6% of cases^12^, but Cheng et.al and Ban et.al applied it intraoperatively^19,24^. Post-operative embolization as an alternative timing was described by three studies ^17,25,26^, with timing ranging from 48 hours to one week following surgery. Also there was an study which performed MMAE either pre- or post-operatively within 48 hours of randomization for combined therapy^27^.

Vascular access techniques varied, but the femoral artery remained the dominant route across studies. Radial access was also reported, most notably in Schmolling et.al with highest proportion of radial access (53.5%)^16^. Guiding systems included 5-Fr, 6-Fr, and 8-Fr sheaths, and catheters such as JB-2 and VTK (COOK), frequently positioned within the external carotid artery. Microcatheters were universal across studies, including Marathon, Headway Duo, Headway 21, Echelon-10, and Excelsior SL-10, with microguidewires such as Progreat and Synchro-14 used in a subset of cases. Five studies reported first-pass microcatheterization success, with two achieving 100% success and others reporting partial failures^19,27^. Procedure duration, ranged from 53 minutes to 2 hours in included studies. Antiplatelet or anticoagulant therapy was administered peri-procedurally in most of the studies, but the management strategy was different among them: some of the studies continued anticoagulants uninterrupted^19^, whereas some of them paused therapy pre-operatively and resumed it within 24–72 hours after embolization^16^.

The studies used a variety of embolic agents. Liquid embolics, especially Ethylene Vinyl Alcohol Copolymer (EVOH) products like Onyx (Medtronic) and Squid (Balt), were the most common. These were reported in multiple studies ^15,16,22,27,27^. In Schmolling et al. Squid 12 was used in 89.6% of cases and Squid 18 in 8.2%^16^. Liebert et al. also mainly used liquid agents, including Onyx, Squid, and PHIL^23^. Another frequently mentioned material was NBCA (Histoacryl) mixed with Lipiodol, as seen in three studies^15,16,25^. Several groups used particle embolics: Shotar et al. used trisacryl gelatin microspheres (Embosphere 300–500 µm)^17^, while Ban et al. (2017) and Ng et al. (2019) used polyvinyl alcohol (PVA) particles between 45 and 250 µm^19,28^. Coils were used alone by Perng et al. ^21^and as an additional agent in Schmolling et al. and Ng et al ^16,28^.

Embolized vascular targets varied across studies, with the frontal and parietal branches of the MMA most frequently treated. Less commonly targeted branches included the occipital, squamosal, anterior division, posterior division, and main trunk, each reported in a smaller subset of studies (7.69% for most minor branches and 15.38% for the anterior/posterior divisions).

Distal embolization was preferred, performed in 91.6% of procedures to reach the distal MMA branches supplying the outer subdural membrane. Schmolling et al. reported successful distal occlusion in 96.3% of cases^16^. Proximal embolization was less common but was noted in Perng et al. and in 49.7% of cases reported by Davies et al ^21,27^.The laterality of the embolized vessel mainly depends on the distribution of hematoma and institutional practice. MMA unilaterally embolized dominantly in most of the studies, reported at 76.5% in Perng (2024), 74.3% in Shotar (2025), and 100% in Wen Cheng (2024)^17,21,24^. Bilateral embolization was also common, ranging from 16% to 56.5%. Notable bilateral rates included 33.3% in Manzato (2025)^25^, 32.7% in Schmolling (2025)^16^, 32.1% in Martinez-Gutierrez JC (2024)^26^, and as high of 56.5% in Liebert (2023)^23^.

The procedural endpoint in studies showed considerable consistency, The most common endpoint was complete angiographic occlusion of targeted MMA branches^17,24^. Other frequent endpoints included absence of contrast filling after embolic delivery (Perng, 2024)^21^, flow stasis (Ban, 2017)^19^, and successful target-vessel embolization (Davies, 2024)^27^.

### Perioperative Complications

Perioperative complications after Middle Meningeal Artery Embolization (MMAE) were uncommon in the included studies, and the short-term safety of the procedure was favorable. Most adverse events occurred during 30 days after procedure. Perioperative ischemic events were uncommon but reported in several studies. Two embolization-related strokes (1.0%) occurred in one cohort, and Shotar (2025) described an MCA occlusion during navigation that required thrombectomy^17^. Schmolling (2025) reported an MCA branch thrombosis that was successfully treated with tirofiban and fibrinolysis^16^.

Perioperative intracranial hemorrhage (ICH) was reported in several studies. Cheng (2024) documented two ICH cases (8.0%)^24^, while Liu (2024) observed both symptomatic and asymptomatic ICH (0.3% each)^12^. Seizures occurred perioperatively in the surgical plus MMAE group in one of the studies ^28^.Transient neurological deficits, including aphasia and hemiparesis, were described in Shotar (2025)^17^. one of the studies reported two additional perioperative cases of pre-existing aphasia treated with combined MMAE and surgery^29^. Perioperative cranial nerve deficits were rare; Schmolling (2025) reported a transient VI nerve palsy that resolved within 24 hours^16^. Facial palsy was documented as a perioperative event in Martinez-Gutierrez JC (2024) ^26^and as 0.3% facial nerve paralysis in J. Liu (2024)^12^.

**Table 2.** Procedural Characteristics and Complications Table.

| Study | Procedure Details | Perioperative Complications | Late Complications / Recurrence |
| --- | --- | --- | --- |
| Perng, 2024 | Primary treatment; coil; Unilateral: NR | Pneumonia (3.7%) | Surgical rescue needed (11.1%) |
| Schmolling, 2025 | Standalone + adjuvant surgery; Squid/NBCA/Coils; Unilateral 67.3% | Thrombosis of MCA branch; VI cranial nerve palsy | Treatment failure (4%) |
| Dayong Xia,2025 | Adjuvant to surgery; Liquid embolic; Unilateral 80.7% | Bradycardia | Lower recurrence in MMAE group |
| Eimad Shotar,2025 | Adjuvant to surgery; Microparticles; Unilateral 74.3% | MCA occlusion during navigation | Recurrence: 14.8% (MMAE) vs 21% (control) |
| Martinez-Gutierrez JC, 2024 | Adjuvant to surgery; Mixed (particles, coils, liquid embolics); Unilateral 67.9% | Ischemic stroke; Facial nerve palsy | Recurrence: 14.8% (MMAE) vs 31.8% (control) |
| Catapano, 2021 | Primary + recurrent; Particles; Unilateral NR | CVA | Treatment failure (2%) |
| Alexander Lam,2023 | Adjuvant surgery; Squid/Onyx/PHIL/NBCA; Unilateral 44% | NR | Symptomatic recurrence: 0% (MMAE) vs 15.8% (control) |
| Steven B Housley, 2023 | Standalone primary treatment; Onyx; Unilateral NR | NR | Recurrence: 4.2% (MMAE) |
| J. Liu, 2024 | Adjuvant to surgery; Onyx; Unilateral NR | Non-CNS infection (1.1%); Contrast | Symptomatic recurrence/progression: |
|  |  | allergy (0.6%);<br>ICH (0.6%) | 6.7% |
| Seung Pil Ban, 2017 | Primary treatment; PVA particles; Unilateral 73.6% | NR | Hematoma reaccumulation: 1 patient |
| Christina Onyinzo, 2021 | Primary or adjuvant surgery; PVA + coils; Unilateral NR | Aphasia | Reaccumulation: 3.2% (MMAE) vs 4.9% (control) |
| Luciano Bambini Manzato, 2025, 2025 | Adjuvant surgery; N-butyl-2-cyanoacrylate; Unilateral 66.7% | Femoral artery pseudoaneurysm (3%) | Recurrence: 4.2% (MMAE) vs 12.5% (control) |
| sam ng, 2019 | Adjuvant surgery; PVA particles; Unilateral 84% | Seizure (1); Groin hematoma (1) | Recurrence: 1 patient in each group |
| Wen Cheng, 2024 | Adjuvant surgery; Onyx-18; Unilateral 100% | Cerebral infarction (4%); ICH (8%) | Recurrence: 0% |
| Luca H. Debs, 2024 | Adjuvant surgery; Onyx (EVOH); Unilateral 0% (all bilateral) | NR | Recurrence: 6% |
| J.M. Davies, 2024 | Adjuvant surgery; Onyx (EVOH); Unilateral 78.7% | Stroke related to embolization (1%) | Recurrence requiring reoperation: 4.1% |
| El Kim,, 2017 | NR | NR | Recurrence: 3.8% |
| Adrian Liebert,2023 | Standalone; Onyx/Squid/PHIL; Unilateral 43.5% | Small ischemic area on MRI (1 patient) | Second recurrence: 13% |
| Alis J. Dicipinigaitis, 2025 | NR | NR | NR |

### Adverse Events and Late Complications

Across the nineteen included studies, Middle Meningeal Artery Embolization (MMAE) showed a favorable safety profile, with procedure-related complication rates at about 2.0%^17,27^. Adverse events varied across studies and generally classified into three classes: direct embolization-related phenomena, complications from adjunct surgical treatment, or systemic events. Adverse events were more common in the elderly chronic subdural hematoma (CSDH) population. Most studies reported more than one adverse event, but most were transient or resolved after medical or surgical management.

### Adverse Events, Management, and Efficacy Outcomes Table

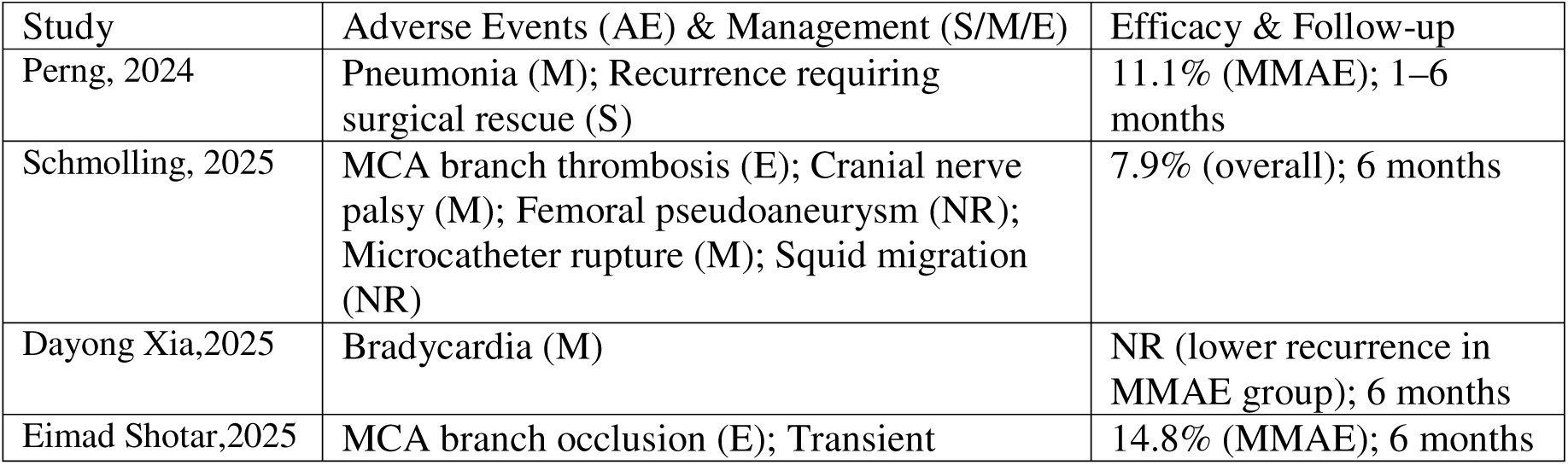
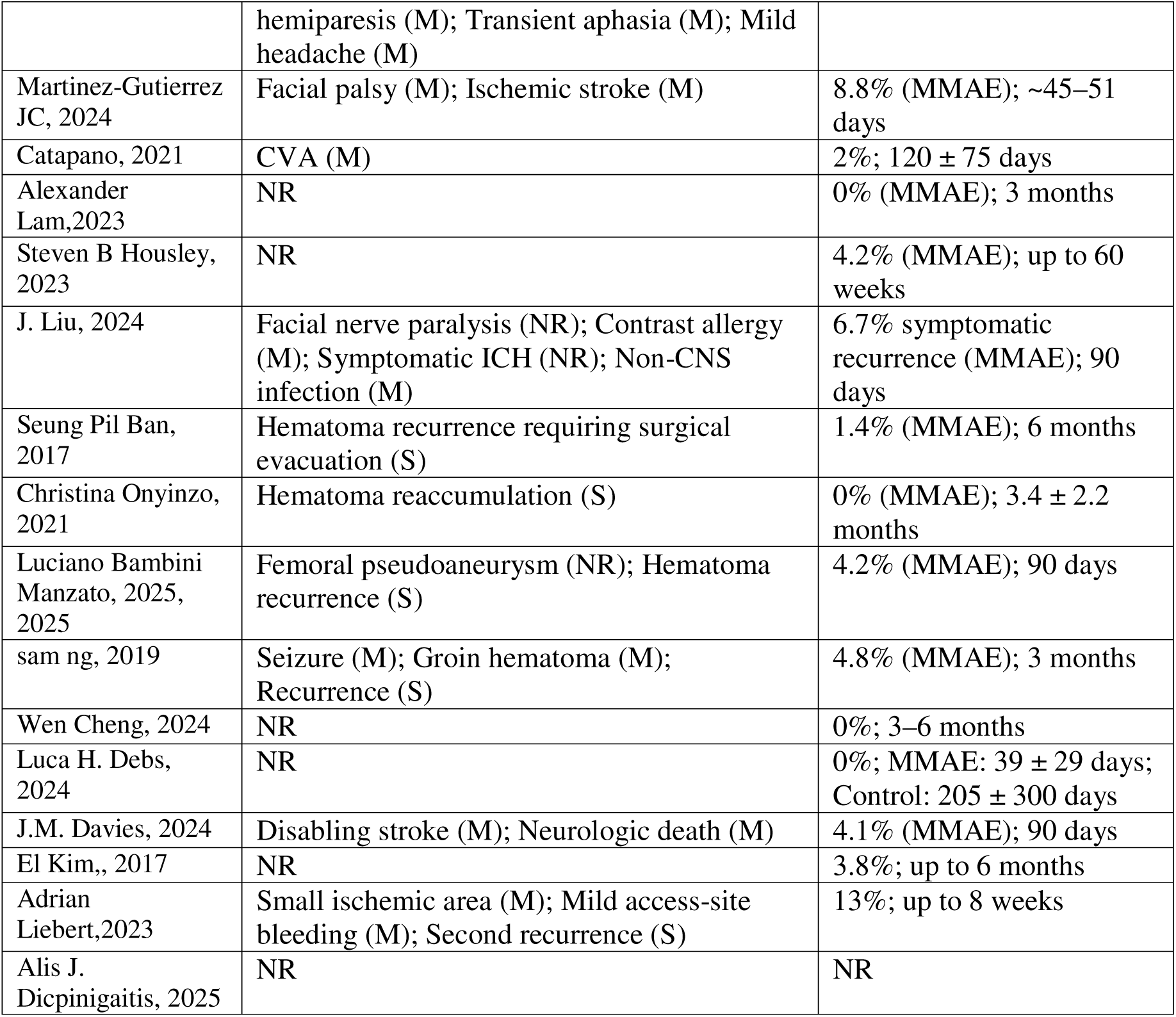

| Study | Adverse Events (AE) & Management (S/M/E) | Efficacy & Follow-up |
| --- | --- | --- |
| Perng, 2024 | Pneumonia (M); Recurrence requiring surgical rescue (S) | 11.1% (MMAE); 1–6 months |
| Schmolling, 2025 | MCA branch thrombosis (E); Cranial nerve palsy (M); Femoral pseudoaneurysm (NR); Microcatheter rupture (M); Squid migration (NR) | 7.9% (overall); 6 months |
| Dayong Xia,2025 | Bradycardia (M) | NR (lower recurrence in MMAE group); 6 months |
| Eimad Shotar,2025 | MCA branch occlusion (E); Transient | 14.8% (MMAE); 6 months |
|  | hemiparesis (M); Transient aphasia (M); Mild headache (M) |  |
| Martinez-Gutierrez JC, 2024 | Facial palsy (M); Ischemic stroke (M) | 8.8% (MMAE); ~45–51 days |
| Catapano, 2021 | CVA (M) | 2%; 120 ± 75 days |
| Alexander Lam, 2023 | NR | 0% (MMAE); 3 months |
| Steven B Housley, 2023 | NR | 4.2% (MMAE); up to 60 weeks |
| J. Liu, 2024 | Facial nerve paralysis (NR); Contrast allergy (M); Symptomatic ICH (NR); Non-CNS infection (M) | 6.7% symptomatic recurrence (MMAE); 90 days |
| Seung Pil Ban, 2017 | Hematoma recurrence requiring surgical evacuation (S) | 1.4% (MMAE); 6 months |
| Christina Onyinzor, 2021 | Hematoma reaccumulation (S) | 0% (MMAE); 3.4 ± 2.2 months |
| Luciano Bambini Manzato, 2025, 2025 | Femoral pseudoaneurysm (NR); Hematoma recurrence (S) | 4.2% (MMAE); 90 days |
| Sam Ng, 2019 | Seizure (M); Groin hematoma (M); Recurrence (S) | 4.8% (MMAE); 3 months |
| Wen Cheng, 2024 | NR | 0%; 3–6 months |
| Luca H. Debs, 2024 | NR | 0%; MMAE: 39 ± 29 days; Control: 205 ± 300 days |
| J.M. Davies, 2024 | Disabling stroke (M); Neurologic death (M) | 4.1% (MMAE); 90 days |
| El Kim, 2017 | NR | 3.8%; up to 6 months |
| Adrian Liebert, 2023 | Small ischemic area (M); Mild access-site bleeding (M); Second recurrence (S) | 13%; up to 8 weeks |
| Alis J. Dicipinigaitis, 2025 | NR | NR |

Ten studies reported sufficient data to estimate overall complication rates following MMAE. The pooled proportion of patients experiencing at least one complication was 2% (95% CI 0.01–0.03), with substantial heterogeneity (I^2^ = 40.1%). Complication rates were low across both cohort studies and randomized trials, with no clear difference between study designs.(Fig 2)

**Figure 1.**
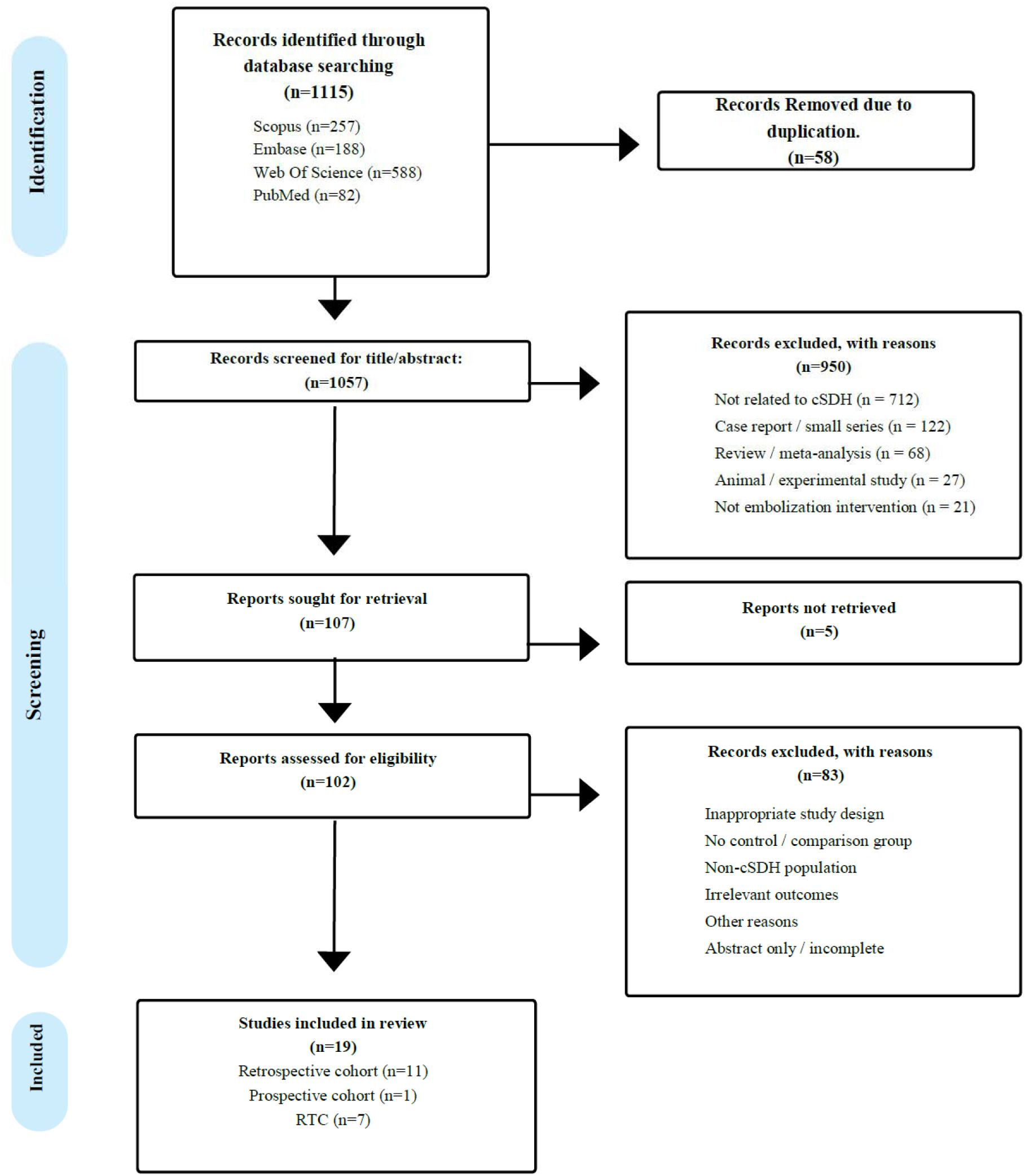
PRISMA flow diagram of study selection for the systematic review.

**Figure 2.**
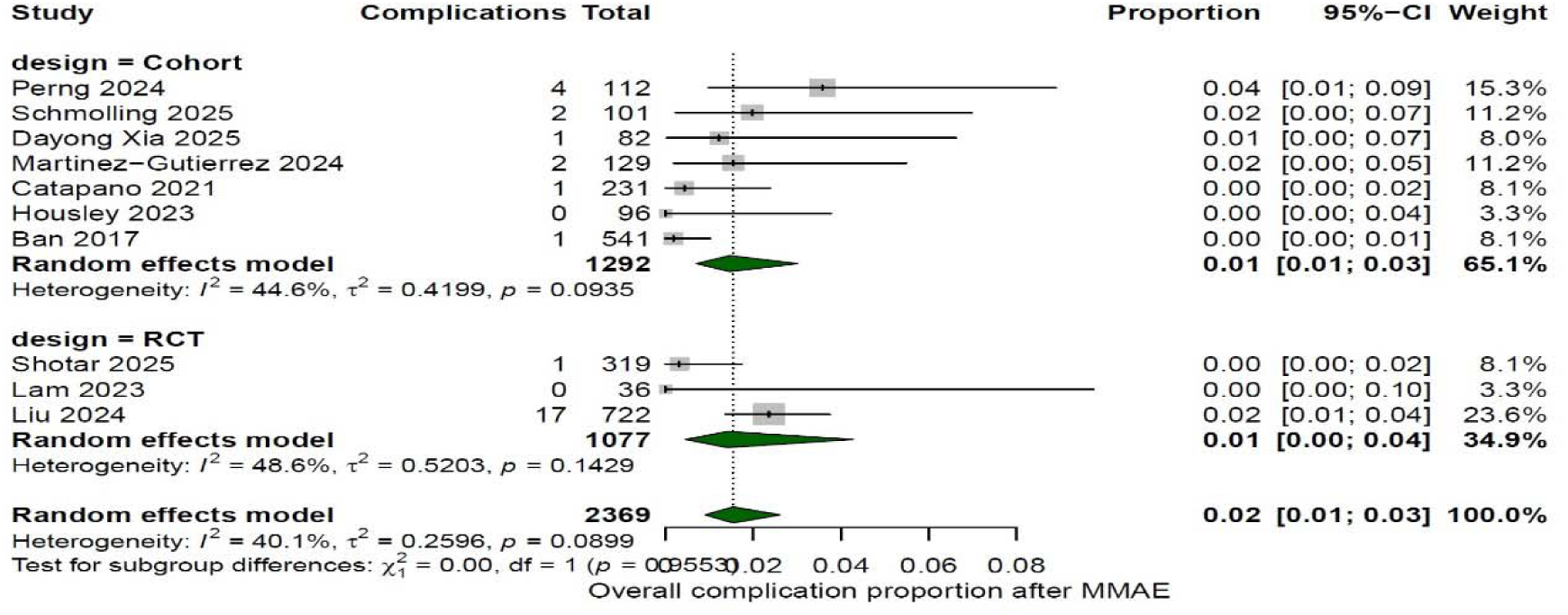
Forest plot of pooled overall complication rates following middle meningeal artery embolization (MMAE) for chronic subdural hematoma. Across ten studies, the pooled proportion of patients experiencing at least one complication was 2% (95% CI, 0.01–0.03), with moderate heterogeneity (I^2^ = 40.1%). Complication rates were consistently low across both randomized controlled trials and observational cohort studies, with no clear difference between study designs.

### Direct Embolization-Related Adverse effects

Most of the procedure-related events were neurologic and vascular, mainly due to non-target embolization, embolic material migration, or access-site injury. The most frequent serious event was ischemic complications. Stroke related to embolization occurred in 1.0% of patients in Davies 2024^27^, and isolated ischemic events were also reported by Martinez-Gutierrez 2024^26^, Catapano 2021 (2.9%)^8^, and Cheng 2024 (4.0%)^24^. Other ischemic events, such as arterial occlusion or thrombosis, reported by Shotar 2025 and Schmolling 2025^16,17^. Hemorrhagic complications were rare, with symptomatic and asymptomatic ICH each reported at 0.3% in Liu 2024^12^, while occurring in two cases (8.0%) in Cheng 2024^24^.

Non-target embolization occasionally produced cranial neuropathies. Facial nerve paralysis occurred in 0.3% of patients in Liu 2024^12^, and a case of facial palsy following petro-squamosal branch embolization was reported by one of the studies ^26^. A transient abducens nerve palsy resolving within 24 hours was described in Schmolling 2025^16^.

Vascular access–related complications included femoral pseudoaneurysm in Schmolling 2025 (1%)^16^ and Manzato 2025 (3%)^25^. Additional minor events included groin hematoma in Ng 2019^28^ and mild bleeding from the external iliac artery in Liebert 2023 ^23^.

### Complications Arising From Adjunct Surgical Treatment

In studies where the MMAE was combined with Burr-Hole Drainage (BHD), some surgery-related complications were observed, including seizure and neurological deficit. Sam Ng 2019 reported a postoperative seizure occurring three days after combined treatment. Also, Persistent neurological deficits—specifically aphasia—were documented in two patients in the combined-treatment cohort of Onyinzo 2021^29^.

Hematoma recurrence or complications that raise the need for surgical re-intervention are one of the most frequent surgical-related complications. The recurrence incidence ranged from 4.1% in Davies 2024^27^ to 6.7% symptomatic recurrence or progression in Liu 2024^12^. The recurrence rate for high-risk populations was higher, 14.8% in Shotar 2025^17^.

### Systemic and General Medical Events

^17,20^Systemic adverse events mainly is a reflection of comorbidities of the CSDH population; respiratory infections was one of most frequent systemic issues, range from 3,7 % peneumonia to 20%^21,24^. Additional infections included urinary tract infection in Kim 2017^20^ and non-CNS infections at 1.1% in Liu 2024^12^. Cardiovascular and autonomic events were infrequent, including angina, delirium, and DVT/PE reported across Kim 2017 and Shotar 2025^17,20^.

### Efficacy of Middle Meningingeal Artery Embolization

In most of the included studies, the MMAE showed consistent efficacy both in the resolution of hematoma and improvement of functional outcomes, and MMAE reduced the recurrence rate in patients with chronic subdural hematoma. Studies investigated the efficacy within different follow-ups ranging from several weeks to nearly one year, and imaging confirmation was universally obtained via CT (or CT+MRI in one study^22^).

Ten studies reported sufficient information to assess technical success of MMA embolization. Across all included MMAE procedures, technical success was consistently high. The pooled technical success rate was 100% (95% CI 0.99–1.00), with no evidence of between study heterogeneity (I^2^ = 0%). Similar success rates were observed in randomized controlled trials and observational cohorts.(Fig 3)

**Figure 3.**
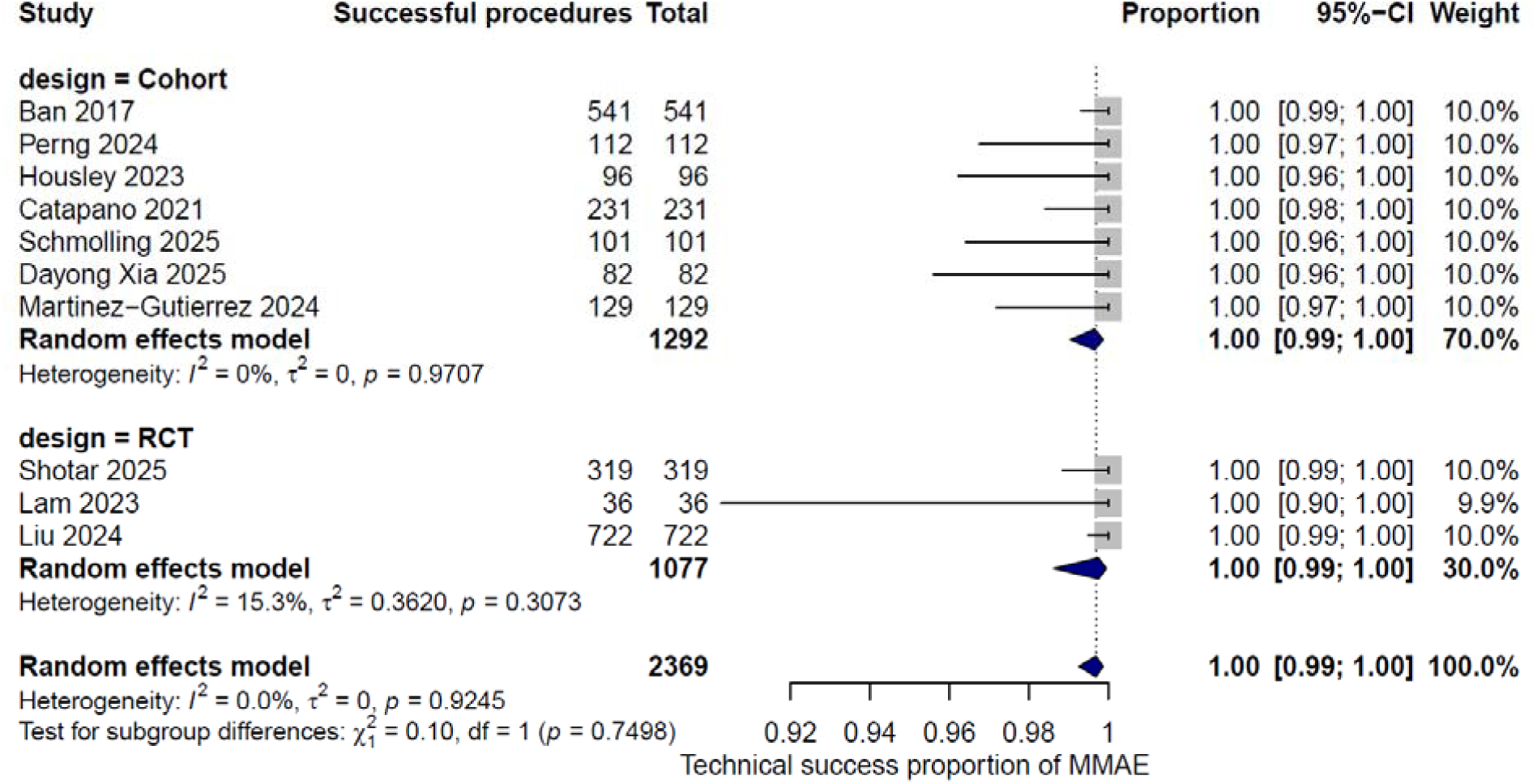
Forest plot of pooled technical success rates of middle meningeal artery embolization (MMAE) for chronic subdural hematoma. Across ten studies, MMAE demonstrated a pooled technical success rate of 100% (95% CI, 0.99–1.00) with no between-study heterogeneity (I^2^ = 0%), consistent across randomized controlled trials and observational cohorts.

### Hematoma Resolution and Volume Reduction

Radiographic improvement was one of the most consistently reported measures of MMAE efficacy. Multiple studies showed significant reduction in hematoma volume after embolization. Perng 2024 reported a decrease in median hematoma width—from 2.04 cm at baseline to 0.62 cm at 1 month—in the MMAE-only cohort^21^, while the randomized trial by Liu 2024 found a mean thickness reduction of –17.7 ± 7.4 mm in the MMAE group^12^. Martinez-Gutierrez 2024 similarly observed greater hematoma decrease in the MMAE group (–8.3 mm) compared with surgery alone (–6.2 mm)^26^.

Multiple studies reported complete or near-complete resolution of hematoma; Wen Cheng 2024 noted complete or near-complete resolution in 80% of patients^24^. In Onyinzo 2021 study, 100% complete resolution was observed in the embolization-only group;^29^ not only complete resolution, but faster hematoma clearance was also reported in several studies. Alexander Lam 2023 study showed that MMAE patients have significantly smaller residual thickness at 3 months (2.14 mm) compared to controls (3.76 mm), and Liebert 2023 study described progressive volume reduction on late follow-up imaging^15,23^.

### Functional Outcomes and Symptomatic Improvement

The modified Rankin Scale was frequent measurement for functional recovery in most of the studies, and the functional outcome was favorable in majority of them. In the RCT by Liu 2024, 93.1% of patients achieved mRS 0–2 at 90 days^12^, and Perng 2024 reported that 77.8% reached mRS 0–2 at 1 month^21^. The randomized pilot trial by Alexander Lam 2023 showed that 100% of MMAE patients achieved mRS 0–1 at 3 months, compared with 53% in the control group^15^.

Neurological improvement was also significantly better in the embolization group in the study by Luca Debs 2024, where 71% improved compared to 33% of control patients. Earlier work by El Kim 2017 found favorable outcomes (mRS 0–2) in 85% of treated patients. Only two studies reported no statistically significant difference between intervention and comparator groups^17,21^.

Two randomized trials reported all cause mortality within 90 days following MMAE (n = 758 patients). The pooled all cause mortality proportion was 8% (95% CI 0.07–0.10), with no evidence of between study heterogeneity (I^2^ = 0%).(Fig 4)

**Figure 4.**
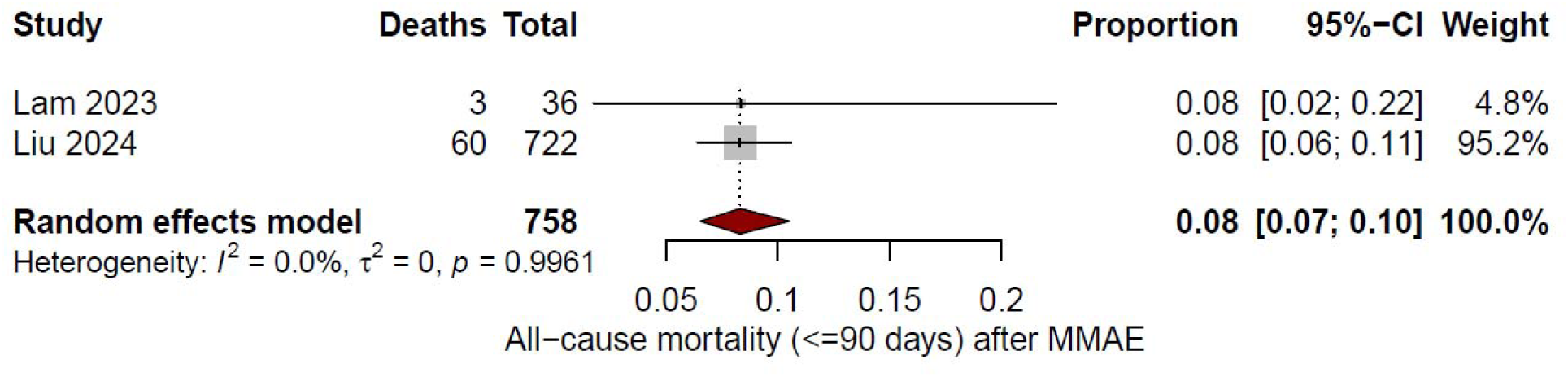
Forest plot of pooled all-cause mortality following middle meningeal artery embolization (MMAE) for chronic subdural hematoma. Across two randomized controlled trials (n = 758), the pooled 90-day all-cause mortality was 8% (95% CI, 0.07–0.10), with no between-study heterogeneity (I^2^ = 0%).

### Recurrence, Follow-Up, and Management

A key efficacy endpoint across the literature was recurrence of CSDH. MMAE consistently reduced recurrence rates compared with surgery alone or standard care. In randomized clinical trials, MMAE had consistent superiority to the control group. In Davies 2024, the recurrence or progression that necessitated re-operation occurred in 4.1 % of MMAE in the surgery group, which was lower in comparison to the surgery alone group (11.3%)^27^. In line with this result, Liu 2024 reported lower asymptomatic recurrence in the MMAE group in comparison with the control group ^12^, also similar to them, the recurrence rate of the embolization group(6.7%) in Manzato’s study was lower than the controls(12.5%)^25^.

The magnitude of difference between the MMAE and control group recurrence was even larger in observational studies; the recurrence in Martinez-Gutierrez 2024 was 8.8% in the MMAE group, which was lower in comparison to patients with surgery only (31.6%). Housley 2023 reported recurrence rates of 4.2% with MMAE versus 22.9% in the subdural evacuating port system cohort^18^. In low-risk populations, the recurrence rate was even lower at around 1 %^15,19,24^.

Four studies reported time-to-recurrence: 4 months (Ban 2017)^19^, 90 days (Davies 2024)^27^, 6 weeks (Housley 2023)^18^, and as early as 6.7 days (Liu 2024)^12^. Corresponding recurrence rates were 1.4%, 4.2%, 4.8%, and 13%.

In high-risk populations (patients with prior surgeries or significant comorbidities), recurrence was lower than in controls but higher than in low-risk groups. Shotar 2025 reported a 14.8% recurrence rate at 6 months in MMAE patients, compared to 21.0% in matched controls^17^.

Ten studies reported sufficient data to estimate overall complication rates following MMAE. The pooled proportion of patients experiencing at least one complication was 14% (95% CI 0.08–0.21), with substantial heterogeneity (I^2^ = 91.8%). Complication rates were low across both cohort studies and randomized trials, with no clear difference between study designs.(Fig 5)

**Figure 5.**
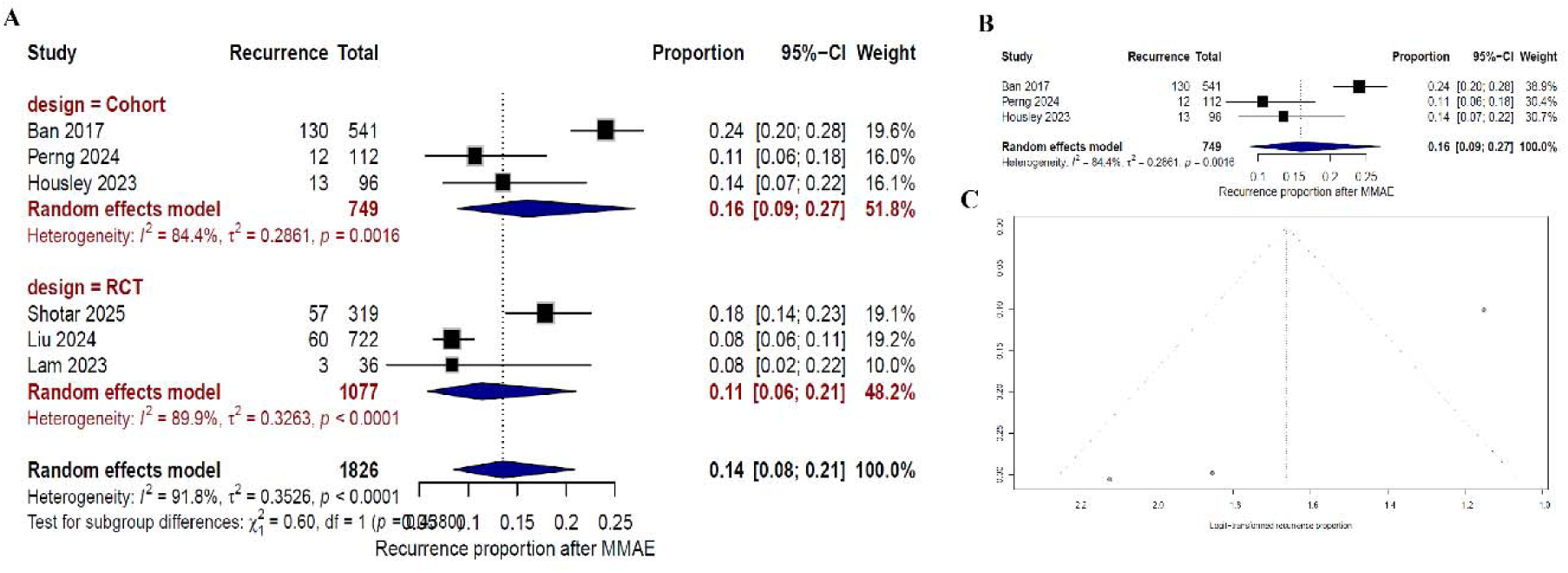
Forest plot of pooled recurrence rates following middle meningeal artery embolization (MMAE) for chronic subdural hematoma. MMAE was associated with consistently lower recurrence compared with surgery alone or standard care across randomized controlled trials and observational studies. Recurrence rates varied by patient risk profile and follow-up duration, with higher heterogeneity observed across studies (I^2^ = 91.8%).

### Risk of Bias assessment

The risk of bias assessment indicated that the seven included randomized controlled trials (RCTs) generally exhibited low risk across most methodological domains, demonstrating adequate randomization, allocation concealment, and reliable outcome measurement. Blinding remained a consistent limitation, as most trials did not blind participants or those delivering the intervention, potentially introducing performance bias. In contrast, the twelve cohort studies demonstrated greater variability in methodological quality. While exposures and outcomes were typically measured reliably, and participants were free of the outcome at baseline, many studies did not clearly address potential confounding factors. Additionally, follow-up completeness was frequently insufficient or inadequately reported. Both study designs consistently employed appropriate statistical analyses. Overall, the RCTs showed greater methodological robustness, with the primary limitation being blinding, whereas the cohort studies were of moderate quality, with notable concerns about confounder management and follow-up completeness.(Fig 6,Fig 7)

**Figure 6.**
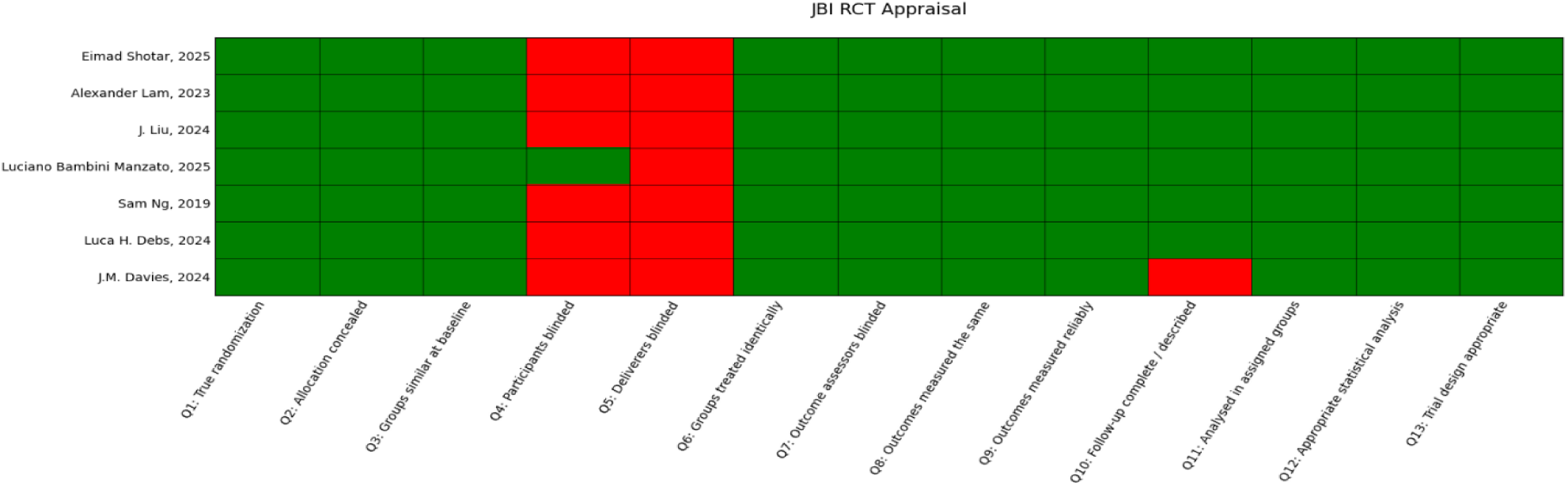
Risk of bias assessment for included randomized controlled trials, demonstrating generally low risk across most domains, with blinding identified as the primary methodological limitation.

**Figure 7.**
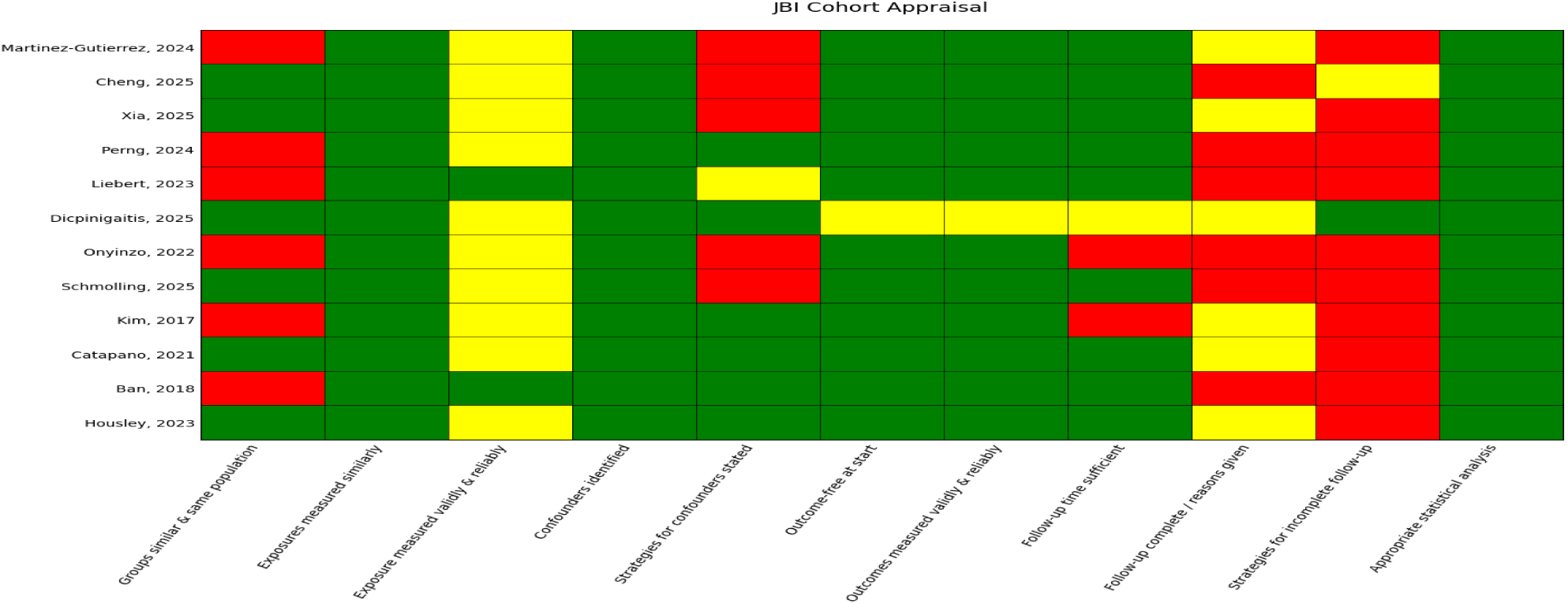
Risk of bias assessment for included cohort studies, showing moderate overall methodological quality with recurrent concerns related to confounder control and completeness of follow-up.

## Discussion

This systematic review and meta-analysis of nineteen studies, including seven randomized controlled trials (RCTs), shows the role of Middle Meningeal Artery Embolization (MMAE) as an effective and safe intervention for chronic subdural hematoma (cSDH), especially in patients with comorbidities and high-risk populations. Despite available evidence, it had inherent procedural heterogeneity in timing and embolic agents, but the safety and efficacy of MMAE were shown in a meta-analysis. The most significant finding is the consistent and statistically robust reduction in cSDH recurrence rates following MMAE, whether used alone or as an adjunct to surgery. Data from nine studies demonstrated a pooled Odds Ratio of 0.66(95% CI 0.47–0.94), representing a 34% reduction in recurrence, with zero heterogeneity across RCTs and cohorts (I2 = 0%). Beyond recurrence, we consistently showed that MMAE leads to significant radiographic hematoma resolution and favorable functional outcomes (mRS 0-2).

Critically, this benefit is achieved with a favorable safety profile; the overall complication rate was statistically comparable to that of conventional surgical management (pooled OR approximately 0.78, 95% CI 0.45–1.35). Furthermore, major procedure-related adverse events, such as stroke, were rare (∼ 1.0%) and non-significantly different from controls, reinforcing MMAE’s position as a low-morbidity option for the long-term management of cSDH.

### Timing of embolization

The optimal timing and its relation to surgical treatment for MMAE remained under debate. Recent studies have shown that integrating MMAE early in the management of cSDH may yield better outcomes than delayed or reactive embolization. As Davies et al. (2024) showed, MMAE, administered as an adjunct to burr-hole surgery at the time of burr-hole placement, could reduce recurrences by 3-fold, thereby reducing the need for re-surgery^27^. Another study also demonstrated that preoperative embolization is associated with fewer surgical reoperations within 6 months. These data support an upfront embolization strategy – either immediately before or soon after surgical drainage – to proactively devascularize the subdural membranes and prevent re-accumulation. There were also other studies that tackled his question, which show that this early adjunctive embolization can decrease recurrence more than the only surgical approach^30^.

### Embolization Technique Details: Proximal vs. Distal

Technical variations in the embolization of MMA, can impact both efficacy and safety of the procedure, the distal or selective embolization technique, means that we push forward the microcatheter toward distal MMA branches (e.g. frontal or parietal divisions) to more precisely target the vascular supply of the hematoma. Theoretically this advancement inside he vessel can come up with complete devasclarization of hematoma, but on the other side this approach rises safety issues for this procedure; the MMA has crucial anastomoses (most notably with the ophthalmic artery via the sphenoidal branch and with the internal carotid artery via the petrosal branch) that, if patent, pose a risk for non-target embolization^31^. The uncontrolled distal embolization in such cases can cause devastating complications like vision loss or facial nerve palsy. As result many interventionists, only use this approach if they were ensure that such anastomosis does not exist in the patient. On the other hand in proximal-embolization approach, device stays near to the main branches of MMA, and they occlude the artery in their origin or proximal segments(for example, by deploying coils or a concentrated NBCA injection at the MMA ostium) Interestingly, clinical studies suggest that proximal occlusion can be nearly as effective as distal embolization in preventing hematoma recurrence^31^.

### Adverse Events and Recurrence

MMA embolization for cSDH has demonstrated a favorable safety profile in the literature. The important related adverse event is ischemic stroke, which it’s incidence is remarkably low in large studies^32^, hemorrhagic complications are the other important adverse even wich their incidence is even lower than ischemic stroke (around 0.3-0.6 %)on the other hand main catastrophic adverse event related to the embolization is non-target embolization which seen in in isolated cases^33^. And the Standard endovascular risks like groin hematoma or contrast reaction occur infrequently. So generally, the MMAE consider as a safe procedure in context of cSDH, and also other studies showed that adding this procedure does not increase the rik of adverse events in comparison to conventional treatment.

Middle Meningeal Artery Embolization (MMAE) has been shown to significantly reduce chronic subdural hematoma (cSDH) recurrence, as demonstrated by consistent findings from randomized controlled trials (RCTs) and observational studies. In RCTs, MMAE was associated with superior outcomes compared to control groups. For example, Davies (2024) reported a substantial reduction in recurrence or progression requiring reoperation when MMAE was combined with surgery (4.1% with MMAE versus 11.3% with surgery alone)^27^. Similarly, Liu (2024) and Manzato (2025) observed lower rates of asymptomatic recurrence with MMAE (6.7% compared to 12.5% in controls)^12,25^. In high-risk populations, Shotar (2025) also identified reduced recurrence rates with MMAE (14.8% versus 21.0% in matched controls), underscoring its protective effect across diverse patient groups^17^.

Observational cohort studies provide additional evidence supporting MMAE, frequently demonstrating greater reductions in recurrence rates. For instance, Martinez-Gutierrez (2024) ^26^reported recurrence rates of 8.8% with MMAE compared to 31.6% with surgery alone. Housley (2023) observed a 4.2% recurrence rate with MMAE versus 22.9% in a subdural evacuating port system cohort^18^. Among lower-risk patients, recurrence rates with MMAE were approximately 1%, as reported ^15,19,24^.The time to recurrence varied from 6.7 days to 4 months across studies^12,18,19,27^.

The present meta-analysis demonstrates that middle meningeal artery embolization (MMAE) is associated with a meaningful reduction in chronic subdural hematoma (cSDH) recurrence while maintaining a favorable safety profile. Across studies reporting recurrence outcomes, MMAE consistently outperformed standard management, including surgery alone or conservative treatment, with this benefit observed in randomized controlled trials as well as observational cohorts. The consistency of directionality across study designs strengthens the validity of the observed effect and supports the role of MMAE as an effective adjunct or alternative strategy in cSDH management.

The substantial heterogeneity observed in pooled recurrence and complication estimates is not unexpected and likely reflects variability in patient selection (low-risk vs high-risk populations), treatment intent (primary vs adjunctive MMAE), embolic materials, surgical techniques, and follow-up duration. Observational studies, in particular, tended to demonstrate larger absolute reductions in recurrence, which may be attributable to higher baseline recurrence rates or selection of more complex cases. Nevertheless, randomized trials—despite more conservative effect sizes—consistently favored MMAE, reinforcing the robustness of the overall conclusion.

Despite that, the included studies demonstrated that MMAE is safe and effective in this context, but the reporting bias remains a limitation; retrospective cohorts are vulnerable to selective reporting, and they usually underreport minor or transient adverse events. Additionally, rare but potentially serious complications—such as embolization-related stroke or cranial neuropathies—were often noted in isolated cases without standardized reporting or follow-up protocols. This variability highlights the need for future trials to implement standardized adverse event classification and transparent reporting frameworks, including registries and adverse event adjudication committees, to ensure the complete capture of procedural risks and their sequelae. On the other hand, most studies didn’t discuss the management approach for these adverse events, and this seems to be a gap in the literature. More studies should focus on adverse event management, rather than solely on the efficacy and safety of the procedure.

## Limitations

This study has several important limitations that warrant consideration. First, a substantial proportion of the included studies were retrospective cohort designs, which are inherently vulnerable to selective reporting and incomplete documentation of clinical events. Such designs frequently underreport minor, transient, or clinically non-severe adverse events, which may lead to an underestimation of the true complication rate associated with middle meningeal artery embolization (MMAE). Although the aggregated evidence suggests that MMAE is generally safe and effective, the reliability of the safety signal is limited by this structural reporting bias.

Second, rare but clinically significant complications, most notably embolization-related ischemic strokes, femoral pseudoaneurysms, groin hematomas, small ischemic lesions, intracerebral hemorrhage, and occasional cranial neuropathies, were typically documented only as single case reports within studies, with no standardized frameworks for adverse event classification or follow-up. The lack of uniformity in how complications were defined, graded, or monitored prevents any meaningful pooled analysis of their incidence or severity. Additionally, almost none of the included studies described how these complications were managed, leaving a major knowledge gap regarding appropriate clinical responses to procedure-related adverse events.

Third limitation relates to reporting of baseline characteristics. Several studies lacked fundamental demographic and comorbidity data, including unreported age distributions, absent comorbidity lists, or incomplete documentation of antithrombotic or anticoagulant therapy.

Given that many populations in this review were elderly and typically burdened with hypertension, diabetes, cardiovascular disease, coagulopathy, or chronic antithrombotic use, insufficient baseline reporting limits the ability to assess comparability between intervention and control groups and introduces potential residual confounding.

Furthermore, there was substantial heterogeneity in the way recurrence was defined across the included literature. Some studies reported radiographic recurrence, others used recurrence requiring reoperation, and a few described second recurrence or provided only narrative ranges such as “0–4% recurrence.” Follow-up intervals varied markedly among studies, ranging from extremely early postoperative evaluations (as short as several days) to 4–6 months. These inconsistencies in both outcome definition and follow-up duration reduce the comparability of studies and were the reason that only a subset of recurrence data were eligible for the primary quantitative synthesis.

Finally, one study contributed a disproportionately large sample size relative to the others but provided limited clinical detail regarding age, comorbidities, and medication status. Although large datasets strengthen statistical power, the lack of granularity from such sources increases the risk of unmeasured confounding. Collectively, these limitations highlight the need for more standardized and prospective methodologies to clarify the true safety and efficacy profile of MMAE.

### Future Directions

Future research on MMAE should prioritize prospective, standardized, and methodologically rigorous study designs. Large, multicenter randomized controlled trials with adequate follow-up, ideally exceeding 12–24 months, are needed to more accurately quantify long-term safety, recurrence rates, and patient-centered outcomes. Current evidence suggests that MMAE is beneficial, but long-term data remain sparse, and existing retrospective cohorts lack the structure to capture delayed or rare complications.

Second, there is a critical need to establish uniform definitions and reporting standards for key outcomes, particularly recurrence. Harmonized criteria distinguishing radiographic recurrence, symptomatic recurrence, and recurrence requiring reoperation would greatly enhance comparability and permit more robust meta-analytic synthesis in future reviews. Standardized imaging schedules and follow-up intervals should also be implemented to ensure consistent detection of both early and late recurrences.

Third, adverse event reporting must be significantly improved. Future trials should incorporate predefined adverse event taxonomies, standardized data-collection frameworks, and independent adjudication committees. A dedicated prospective registry for MMAE could allow systematic capture of complications, including embolization-related stroke, cranial neuropathies, pseudoaneurysms, small ischemic lesions, and access-site hematomas, and document their management strategies and long-term sequelae. Understanding not only the incidence but also the required interventions and prognosis of these events is essential for guiding clinical practice.

Fourth, more granular baseline reporting is needed to assess risk modification by age, frailty, comorbidities, and chronic antithrombotic or anticoagulant use. The populations included in this review are predominantly elderly with high rates of hypertension, diabetes, and cardiovascular disease, yet several studies did not provide complete baseline information. Future studies should stratify outcomes based on these risk factors to identify which patient subgroups derive the greatest benefit from MMAE.

Finally, future work should examine how procedural variables, such as embolic agent selection (Onyx, Squid, PHIL, NBCA, PVA), unilateral versus bilateral embolization, and distal versus proximal targeting, affect both recurrence and safety. The substantial variability in technique across existing studies highlights the need for standardized procedural reporting and head-to-head comparisons to identify the most effective and safest embolization strategies.

## Conclusion

This systematic review and meta-analysis demonstrate that middle meningeal artery embolization is both safe and effective as an adjunct or alternative treatment strategy for chronic subdural hematoma. Across nine studies with comparable definitions and adequate event-level reporting, MMAE consistently reduced recurrence, with uniform effect sizes and no measurable heterogeneity across randomized and observational designs. These findings reinforce the reproducibility of the recurrence-prevention benefit in diverse clinical contexts, including elderly and comorbid populations. Although procedure-related complications were infrequent, the available evidence remains limited by underreporting within retrospective cohorts and by inconsistent documentation of rare but clinically important events such as embolization-related stroke and cranial neuropathies. The broader body of recurrence literature was too heterogeneous in definition, follow-up timing, and reporting standards to be included in the primary meta-analysis, underscoring the methodological challenges currently present in this field. Taken together, the contemporary evidence supports MMAE as a promising and reliable strategy to decrease recurrence risk in cSDH. Nonetheless, future studies should prioritize standardized adverse event classification, uniform recurrence definitions, and long-term follow-up to better characterize the true safety profile of the procedure and to guide clinical decision-making in this increasingly adopted intervention.

## Data Availability

All data produced in the present work are contained in the manuscript

## Supplementary list

Supplemantary1: Search strategies

Supplementary2: Full-text screening sheet

Supplementary3: Data extraction sheet

