## Supplementary material for "Efficacy and Safety of Middle Meningeal Artery Embolization in Chronic Subdural Hematoma: A Comprehensive Systematic Review and Meta-Analysis": supp_1

| 12 sep 2025 | | |
| --- | --- | --- |
| **Pubmed** | ("meningeal Artery" OR "meningeal arteries" OR MMA OR MMAE OR transarterial) AND (embolotherapy OR embolotherapies OR "therapeutic embolization*" OR "therapeutic embolisation*" OR "artificial emboli∗" OR "therapeutic occlusion*" OR embol∗ OR endo∗ OR intervention*) AND ("chronic subdural hematoma∗" OR "chronic subdural hemorrhage∗" OR "chronic subdural hemorrhage∗" OR "refractory subdural hematoma∗" OR "refractory subdural hematoma∗" OR "refractory subdural hemorrhage∗" OR "refractory subdural hemorrhage∗" OR cSDH OR "refractory subdural" OR "recurrent subdural" OR "refractory chronic subdural" OR "recurrent chronic subdural" OR "subdural hematoma" OR "subdural bleeding") | 599 |
| **Scopus** | TITLE-ABS-KEY ( ( "meningeal artery" OR "meningeal arteries" OR "middle meningeal artery" OR MMA OR MMAE OR transarterial ) AND ( embolotherapy OR embolotherapies OR "therapeutic embolization*" OR "therapeutic embolisation*" OR "artificial emboli*" OR "therapeutic occlusion*" OR embol* OR endo* OR intervention* ) AND ( "chronic subdural hematoma*" OR "chronic subdural haemorrhage*" OR "chronic subdural hemorrhage*" OR "refractory subdural hematoma*" OR "refractory subdural haemorrhage*" OR "refractory subdural hemorrhage*" OR cSDH OR "refractory subdural" OR "recurrent subdural" OR "refractory chronic subdural" OR "recurrent chronic subdural" OR "subdural hematoma" OR "subdural haemorrhage" OR "subdural hemorrhage" OR "subdural bleeding" ) ) | 677 |
| **WOS** | TS=(("meningeal artery" OR "meningeal arteries" OR "middle meningeal artery" OR MMA OR MMAE OR transarterial)  AND  (embolotherapy OR embolotherapies OR "therapeutic embolization*" OR "therapeutic embolisation*" OR "artificial emboli*" OR "therapeutic occlusion*" OR embol* OR endo* OR intervention*)  AND  ("chronic subdural hematoma*" OR "chronic subdural haemorrhage*" OR "chronic subdural hemorrhage*" OR "refractory subdural hematoma*" OR "refractory subdural haemorrhage*" OR "refractory subdural hemorrhage*" OR cSDH OR "refractory subdural" OR "recurrent subdural" OR "refractory chronic subdural" OR "recurrent chronic subdural" OR "subdural hematoma" OR "subdural haemorrhage" OR "subdural hemorrhage" OR "subdural bleeding")) | 594 |
| **Cochrane trials** | #1 ("meningeal artery":ti,ab,kw OR "meningeal arteries":ti,ab,kw OR "middle NEXT meningeal NEXT arter*":ti,ab,kw OR MMA:ti,ab,kw OR MMAE:ti,ab,kw OR transarterial:ti,ab,kw)  #2 (embolotherapy:ti,ab,kw OR embolotherapies:ti,ab,kw OR (therapeutic NEXT embolization*):ti,ab,kw OR (therapeutic NEXT embolisation*):ti,ab,kw OR (artificial NEXT emboli*):ti,ab,kw OR (therapeutic NEXT occlusion*):ti,ab,kw OR embol*:ti,ab,kw OR endo*:ti,ab,kw OR intervention*:ti,ab,kw)  #3 ((chronic NEXT subdural NEXT hematoma*):ti,ab,kw OR (chronic NEXT subdural NEXT haemorrhage*):ti,ab,kw OR (chronic NEXT subdural NEXT hemorrhage*):ti,ab,kw OR (refractory NEXT subdural NEXT hematoma*):ti,ab,kw OR (refractory NEXT subdural NEXT haemorrhage*):ti,ab,kw OR (refractory NEXT subdural NEXT hemorrhage*):ti,ab,kw OR cSDH:ti,ab,kw OR "refractory subdural":ti,ab,kw OR "recurrent subdural":ti,ab,kw OR "refractory chronic subdural":ti,ab,kw OR "recurrent chronic subdural":ti,ab,kw OR "subdural hematoma":ti,ab,kw OR "subdural haemorrhage":ti,ab,kw OR "subdural hemorrhage":ti,ab,kw OR "subdural bleeding":ti,ab,kw)  #4 #1 AND #2 AND #3 | 65 |
| **Embase** | ('meningeal artery':ti,ab,kw OR 'meningeal arteries':ti,ab,kw OR 'middle meningeal artery':ti,ab,kw OR mma:ti,ab,kw OR mmae:ti,ab,kw OR transarterial:ti,ab,kw) AND (embolotherapy:ti,ab,kw OR embolotherapies:ti,ab,kw OR 'therapeutic embolization*':ti,ab,kw OR 'therapeutic embolisation*':ti,ab,kw OR 'artificial emboli*':ti,ab,kw OR 'therapeutic occlusion*':ti,ab,kw OR embol*:ti,ab,kw OR endo*:ti,ab,kw OR intervention*:ti,ab,kw) AND ('chronic subdural hematoma*':ti,ab,kw OR 'chronic subdural haemorrhage*':ti,ab,kw OR 'chronic subdural hemorrhage*':ti,ab,kw OR 'refractory subdural hematoma*':ti,ab,kw OR 'refractory subdural haemorrhage*':ti,ab,kw OR 'refractory subdural hemorrhage*':ti,ab,kw OR csdh:ti,ab,kw OR 'refractory subdural':ti,ab,kw OR 'recurrent subdural':ti,ab,kw OR 'refractory chronic subdural':ti,ab,kw OR 'recurrent chronic subdural':ti,ab,kw OR 'subdural hematoma':ti,ab,kw OR 'subdural haemorrhage':ti,ab,kw OR 'subdural hemorrhage':ti,ab,kw OR 'subdural bleeding':ti,ab,kw) | 750 |
| **Duplicate** | | 1505 |
| **Final screening** | |  |
